# Sex differences in personal and work-related factors associated with impaired mental well-being among Swiss General Internal Medicine physicians

**DOI:** 10.1101/2025.08.27.25334568

**Authors:** Isaac Egger, Christa Nater, Sven Streit, Jeanne Moor

## Abstract

**Introduction and aims:** Physicians are at risk for impaired mental health. In Switzerland, a lack of understanding of personal and workplace-related factors contributing to impaired physician well-being in women and men hinders the design of effective strategies to improve mental health of the healthcare workforce. In the present work, we focused on physicians of the General Internal Medicine (GIM) workforce and aimed to sex-specifically assess factors associated with a pathological physician well-being index.

**Methods:** We performed a cross-sectional study using an online survey distributed by the Swiss GIM physician society’s newsletter, a journal advertisement, and by clinics. Participants completed the survey between December 2021 and April 2022 and responded to questionnaires assessing personal and workplace-related topics as well as demographic data. The main outcome was the physician well-being index used as a binary variable (pathological vs. normal), and the exposures of interest were different personal and workplace-related factors that may be important for men or women among the GIM physician workforce. We used Chi-squared test to determine sex differences among the physicians with impaired well-being, and we established univariable and multivariable logistic regression models in the whole population of participants and in sex strata order to determine factors associated with impaired physician well-being.

**Results:** This analysis comprised 682 physicians, among them 278 (41%) men and 404 (59%) women. Participants’ mean age was 37 ± 11 years (mean ± standard deviation [SD]). Most participants lived in German-speaking Switzerland (n=485, 71%) and worked in non-academic hospitals (n=267, 39%) and academic hospitals (n=257, 38%). Overall, 18% showed an impaired mental well-being, with little evidence for a sex difference (15% among women vs. 19% among men; p=0.18). Logistic regression analyses revealed some associations between physician well-being and personal factors such as residing in the German-speaking region of Switzerland. Interesting sex-specific associations with parameters of workplace inclusiveness emerged. In fact, sex-stratified analyses revealed that men’s better well-being was associated with greater support by their spouse, whereas women’s well-being is vulnerable to experiences of gender-related discrimination as well as workplace-related conflicts regarding pregnancy and motherhood.

**Conclusion:** Overall, the present data revealed that mostly workplace-related factors are associated with impaired physician well-being, particularly in women. For men physicians, personal factors such as the support by their partners was key. Knowledge on the impact that workplace parameters have on well-being is the critical first step that can inform strategies aimed at improving equal opportunities for mental health and career success among women and men in General Internal Medicine and to ultimately help prevent attrition from the healthcare workforce.

## Introduction

Physicians routinely assess, diagnose and treat diseases of their patients. However, physicians can also present a bad example for their patients due to well-recognized tendencies to disregard their own health (1). Electronic medical records and increasingly complex reimbursement schemes require extensive documentation for optimized billing. This contributes to the problem that physicians-in-training are left with only around 25-30% of their time directly spent on patient care (2,3). With increasing administrative workloads and decreasing autonomy on the job, pressure on physicians rises and causes psychosocial stress and impaired well-being (4,5).

Physician well-being has become a topic of interest following studies among U.S. healthcare professionals. While generally the most widely used tool to assess burnout has historically been the Maslach Burnout Inventory (6), for the case of healthcare professionals a so-called “well-being index” (WBI) has been developed by Dyrbye et al., initially to screen medical students for low mental well-being, burnout and fatigue (7,8). The WBI consisting of 7 questions was then validated in large studies in U.S. physicians (9,10) as part of the Mayo Clinic Program on Physician Well-being and has been named physician well-being index (PWBI).

In surveys on mental well-being conducted among Swiss physicians during the last 20 years, participants working in GIM were predominantly men due to the demographics of the workforce at the time (4). Back then, participants showed moderate but advancing degrees of burnout symptoms over time, most strongly in general practitioners (GPs) (4). However, many of these studies do not reflect the current physician workforce well with its increasing proportion of women among the physician workforce. Data among current and future GPs in Switzerland emerging in the last years showed that well-being was lower in women than in men (11). GIM residents of higher age and not having children were more likely to have an impaired well-being (5). Overall, the potentially modifiable personal and workplace-related factors associated with impaired mental health in Swiss physicians remain an important gap in knowledge. These factors deserve investigation and could serve as a starting point for strategies aiming at improving physician well-being.

Aim of the present study was to investigate the sex-specific determinants of impaired well-being among physicians working in GIM in Switzerland, one of the largest specialties in medicine in Switzerland. To this end, we used a well-validated physician well-being assessment tool in a cross-sectional survey study. Our objectives were to determine the factors associated with impaired PWBI among physicians in GIM in Switzerland, and to investigate potential sex differences in PWBI and in factors associated with PWBI. We hypothesized that both personal and workplace-related factors are associated with impaired mental well-being among GIM physicians in Switzerland, and that these associations underlie sex differences.

## Materials and methods

### Design, setting and participants

We analyzed data obtained through a one-time cross-sectional survey study designed to primarily assess sex differences of well-being and career ambitions among physicians. Data on career ambitions have previously been published (12). The study population were physicians working in GIM in Switzerland. The sole exclusion criterium for the present analysis was non-binary sex because sex-specific subgroup analyses might reveal the identities of the few individuals enrolled in the study with self-reported non-binary sex.

We recruited study participants via the GIM departments of 14 Swiss hospitals, 6 ambulatory clinics or primary care institutes, via an unpaid advertisement in a newsletter by the Swiss Society of General Internal Medicine, via an e-mail by the Swiss Young General Practitioners Association (JHaS), and by a written advertisement in the Swiss medical journal Primary Hospital Care. The anonymous participants were aware of the study’s purpose and gave informed consent. Outcome of the present analysis was a pathological “physician well-being index” (PWBI) (9,10). As a secondary analysis, we determined the sex difference in the overall number of responses of the PWBI that were answered with “Yes”. For the sample size, we aimed for 80% power and a level of significance of 5%, using a web-based tool at https://www.stat.ubc.ca/~rollin/stats/ssize/ to enable the detection of sex differences in impaired mental health. We assumed a prevalence of individuals with pathological PWBI score ≥5 in 20% based on the literature (9). Hence, we determined that to detect a 10% difference in PWBI prevalence between sexes would require n=250 participants of each sex.

### Tools

From December 2021 to April 2022, the study survey was available on the website www.Surveymonkey.com, covering a maximum of up to 109 close-ended or open-ended questions. For the present analysis, we considered only close-ended questions, and most answers were chosen via radio buttons. The questions covered the following topics: First, demographic information was collected including respondents’ age, language region in Switzerland, type of workplace and hierarchical position. Second, mental well-being was assessed by the seven-item PWBI (9,10). A cut-off of ≥5 out of 7 PWBI points was considered impaired mental well-being. The survey also assessed factors known to influence career ambitions such as having a good network and mentoring (14). We determined job satisfaction with a 5-point Likert question («How satisfied are you with your job?»). Workplace environment was assessed using four close-ended questions selected from a 19- item questionnaire of Hall et al. (15), focusing on gender-related and general corporate inclusiveness. Finally, family-related topics identified from the literature (16) were covered including the intent to delay to have children, age at birth of first child, number of children, having adequate childcare, childcare duties and household duties compared with partner, and how parenthood is viewed at workplace. Some pregnancy-related questions were only asked when female participants reported to have been pregnant in the past. Next, the survey contained 8 questions to assess gender discrimination. Because of the unavailability of dedicated tools for physicians, we developed these questions with inspiration by a web-based tool (17) and the literature (18–20).

An initial test version of the survey was provided to 10 physician colleagues to verify comprehensiveness and correct links between survey parts. The final version of the survey underwent back-and-forth translation from English to German or French and back by native speakers to ensure validity (21), and it was available as a German and French version.

### Ethics

The ethics committee of the canton of Bern exempted this study from full review (study ID: Req-2021-01085).

### Analyses

We reported continuous data as mean (standard deviation) or median (interquartile range) as indicated, and categorical data as n (%). For regression models, we categorized all answers from 5-point or 7-point Likert scales as binary values, with the intent to have a simplified overview of “normal” versus “abnormal”. We first performed descriptive statistics, assessing the numbers of answer options and numeric distributions including minimum and maximum of each response value, to identify unreliable answers. Next, we performed Chi-squared and Wilcoxon rank sum test to assess sex differences in the fraction of individuals with a pathological PWBI, and the sex difference in overall PWBI scores, respectively. We next established univariable models of personal and workplace-related factors as exposures of interest in association with binary PWBI (“pathological” vs. “normal”).

To build multivariable regression models, we first chose age, type of workplace and Swiss language region as *a priori* covariates, and we used sex either as a covariate or as stratification variable, as indicated. From the results of univariable models, we aimed to use the variables associated with PWBI for inclusion as potential confounders in multivariable models. However, all identified associations between different factors and PWBI, we determined that these factors may not act merely as confounders but rather act on a common causal pathway. For example, having a good network, receiving one’s supervisor’s support of one’s career, and having good mentors may all be causally linked in their association with mental health, assessed by PWBI. Therefore, we decided to restrict multivariable regression models to the core set of *a priori* covariates that are unlikely to lie on a common causal pathway. Consequently, models were adjusted for age, type of workplace, Swiss language region and sex, unless sex was used as stratification variable wherever indicated. We performed a complete-case analysis. We assessed interactions by comparing associations between individual variables and PWBI between the two sex strata. We performed the analyses with RStudio and considered p-values <0.05 as significant without adjustment for multiple hypothesis testing.

## Results

### Population characteristics

The survey was completed by 684 individuals. We excluded two respondents (0.3%) who reported to have nonbinary gender from analyses, to preserve confidentiality. The final dataset thus comprised 682 individuals. Demographic data of the study population is shown in Table 1. Overall, the population contained 278 (41%) men and 404 (59%) women. Mean age was 37 ± 11 years (mean ± standard deviation [SD]). Most participants were from the German-speaking language region of Switzerland, with n=485 (71%) individuals, followed by the French-speaking region (n=184, 27%) and the Italian-speaking region (n=13, 1.9%). Participants were more likely to work in non-academic hospitals (n=267, 39%) or academic hospitals (n=257, 38%), than in private practice.

**Table 1.** Physician characteristics.

| **Table 1. Physician characteristics** |  |  |  |
| --- | --- | --- | --- |
| **Characteristic** | **N** | **Men,**<br>**N=278** | **Women,**<br>**N=404** |
| Age (years) | 682 | 39 (12) | 36 (10) |
| Swiss language region | 682 |  |  |
| German-speaking |  | 197 (71%) | 288 (71%) |
| French-speaking |  | 73 (26%) | 111 (27%) |
| Italian-speaking |  | 8 (2.9%) | 5 (1.2%) |
| Type of workplace | 682 |  |  |
| University hospital |  | 104 (37%) | 155 (38%) |
| Other place |  | 7 (2.5%) | 18 (4.5%) |
| Other hospital |  | 115 (41%) | 152 (38%) |
| Private practice with <3 colleagues |  | 29 (10%) | 36 (8.9%) |
| Private practice with ≥3 colleagues |  | 23 (8.3%) | 43 (11%) |
| Current work quota (%) |  | 95 (14) | 87 (19) |
| Unknown |  | 37 | 49 |

Data displayed as n (%) or mean (standard deviation)

### Sex differences in prevalence of impaired mental health

Impaired mental health in participating physicians was assessed by the PWBI (Table 2). A bivariate analysis of PWBI and sex revealed some evidence that the fraction with impaired PWBI tended to be smaller among men with 40/262 (15%) than among women with 76/392 (19%), yielding a Chi-Squared test p=0.18. There was stronger evidence for a sex difference in PWBI assessed by the secondary analysis strategy, using the raw numeric PWBI scores, with a median of 2 (IQR: 1-4) of the 7 questions answered “yes” in men (mean ± SD) and 3 (IQR: 1-4) in women (p<0.001). Overall, we found some evidence that impaired physician well-being was more frequent among women than men in the present dataset.

**Table 2.** Sex differences in physician well-being.

Data displayed as n (%) or mean (standard deviation)
| **Table 2. Sex differences in physician well-being** |  |  |  |  |
| --- | --- | --- | --- | --- |
| **Characteristic** | **N** | **Men,**<br>**N=278** | **Women,**<br>**N=404** | **p-value** |
| **Physician well-being index** | 654 |  |  | 0.176 |
| normal |  | 222 (85%) | 316 (81%) |  |
| pathological |  | 40 (15%) | 76 (19%) |  |
| Unknown |  | 16 | 12 |  |
| **Number of abnormal (“Yes”) responses in physician well-being index** | 654 | 2 (1; 4) | 3 (1; 4) | **<0.001** |
| Unknown |  | 16 | 12 |  |

Data displayed as n (%) or median (interquartile range). Physician well-being index contains 7 questions, with threshold for abnormal $\geq 5$ . Statistical analysis by Chi-square or Wilcoxon rank sum test.

### Factors associated with impaired mental health among all physicians

As a next step, we aimed to determine factors that are associated with PWBI among all participants. In univariable analyses, we found that type of workplace, Swiss language region, measures of mentorship, and several measures of workplace inclusivity or gender-related discrimination were associated with PWBI (Supplemental Table 1).

In a multivariable model containing all *a priori* covariates sex, age, type of workplace and Swiss language regions, working in the French-speaking language region was associated with a higher probability of impaired PWBI, with an Odds Ratio (OR) of 1.85 (95% confidence interval [CI]: 1.17, 2.91) (Table 3). Non-academic hospitals and other workplaces were associated with a lower probability of impaired PWBI. There was some weak evidence that female sex was associated with an impaired PWBI, with OR 1.4 (95% CI: 0.91, 2.18, p=0.13).

**Table 3:**
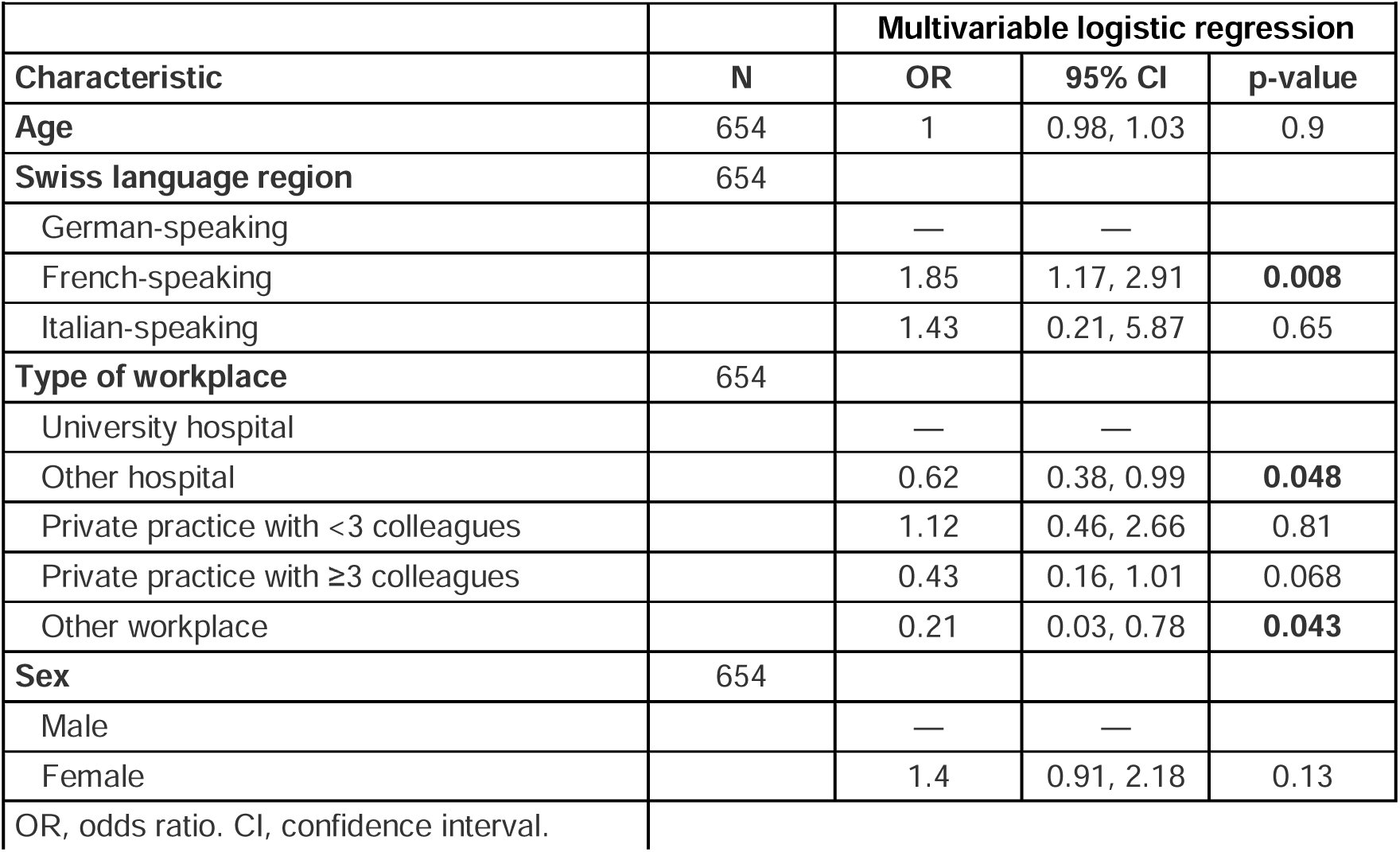
A multivariable logistic regression model of all *a priori* factors associated with impaired physician well-being index.

|  |  |  |  |  |
| --- | --- | --- | --- | --- |
|  |  | **Multivariable logistic regression** |  |  |
| **Characteristic** | **N** | **OR** | **95% CI** | **p-value** |
| **Age** | 654 | 1 | 0.98, 1.03 | 0.9 |
| **Swiss language region** | 654 |  |  |  |
| German-speaking |  | — | — |  |
| French-speaking |  | 1.85 | 1.17, 2.91 | **0.008** |
| Italian-speaking |  | 1.43 | 0.21, 5.87 | 0.65 |
| **Type of workplace** | 654 |  |  |  |
| University hospital |  | — | — |  |
| Other hospital |  | 0.62 | 0.38, 0.99 | **0.048** |
| Private practice with <3 colleagues |  | 1.12 | 0.46, 2.66 | 0.81 |
| Private practice with ≥3 colleagues |  | 0.43 | 0.16, 1.01 | 0.068 |
| Other workplace |  | 0.21 | 0.03, 0.78 | **0.043** |
| **Sex** | 654 |  |  |  |
| Male |  | — | — |  |
| Female |  | 1.4 | 0.91, 2.18 | 0.13 |
| OR, odds ratio. CI, confidence interval. |  |  |  |  |

In subsequent analyses, we adjusted multivariable models for the same core set of 4 variables but added one additional exposure of interest each time (Table 4). In these analyses, aiming for a leading position was associated with a lower probability of impaired PWBI. However, being unhappy with work, reporting an impaired workplace inclusiveness or experiencing gender-related discrimination were associated with a higher probability of impaired PWBI. There was additionally some weak level of evidence that having delayed or planning to delay having children, sensing a negative view of one’s parenthood at the workplace and a lack of one’s partner’s support for the career were associated with a higher probability of impaired PWBI. Overall, the present analyses revealed that several personal but also workplace-related factors were associated with an altered probability of having impaired mental well-being assessed by PWBI among physicians in GIM in Switzerland.

**Table 4:** Multivariable logistic models for personal and workplace-related factors associated with having an impaired physician well-being index.

|  |  |  |  |  |
| --- | --- | --- | --- | --- |
| **Characteristic** | **N** | **OR<sup>1</sup>** | **95% CI<sup>1</sup>** | **p-value** |
| Having a good network: Yes (vs ref.: No) | 586 | 0.37 | 0.23, 0.59 | **<0.001** |
| Having good mentoring: Yes (vs ref.: No) | 586 | 0.37 | 0.22, 0.58 | **<0.001** |
| Being unhappy with work: Yes (vs ref.: No) | 608 | 8.29 | 5.01, 13.9 | **<0.001** |
| My workplace has a culture valuing contributions of both women and men: No (vs ref.: Neutral+Yes) | 630 | 1.68 | 0.92, 2.99 | 0.082 |
| At my workplace, people of different origin are respected: No (vs ref.: Neutral+Yes) | 630 | 1.49 | 0.56, 3.56 | 0.39 |
| At my workplace, a safe environment is promoted in which different people can express their true self: No (vs ref.: Neutral+Yes) | 630 | 2.16 | 1.33, 3.48 | **0.002** |
| Interactions between people of different groups (e.g gender, ethnicity, age) are trustful and respectful at my workplace: No (vs ref.: Neutral+Yes) | 630 | 1.79 | 0.77, 3.90 | 0.15 |
| Delaying having children (ref.: Have not delayed) | 485 |  |  |  |
| have delayed |  | 2.01 | 0.98, 4.11 | 0.054 |
| will delay |  | 2.01 | 0.94, 4.38 | 0.075 |
| will not delay |  | 0.8 | 0.31, 1.96 | 0.63 |
| Number of children (per child) | 654 | 0.95 | 0.75, 1.19 | 0.65 |
| How parenthood is viewed at the workplace: Negative (ref.: Neutral+Good) | 225 | 3.01 | 0.83, 9.99 | 0.078 |
| Having partner's support for professional advancement: No (vs ref.: Neutral+Yes) | 511 | 2.39 | 0.80, 6.41 | 0.094 |
| **Gender discrimination by... / Because of my gender ...** |  |  |  |  |
| Not being taken seriously: Yes (vs ref.: Neutral+No) | 618 | 1.62 | 0.96, 2.72 | 0.066 |
| Others having less confidence in me: Yes (vs ref.: Neutral+No) | 618 | 1.19 | 0.68, 2.03 | 0.54 |
| Feeling discriminated at work: Yes (vs ref.: Neutral+No) | 618 | 2.16 | 1.30, 3.58 | **0.003** |
| Being treated worse by colleagues: Yes (vs ref.: Neutral+No) | 618 | 3.62 | 2.10, 6.19 | **<0.001** |
| Smaller perceived chance to rise in hierarchy: Yes (ref.: Neutral+No) | 618 | 1.59 | 0.94, 2.67 | 0.083 |
| Often taking on duties that don't advance one's career: Yes (vs ref.: Neutral+No) | 618 | 2.06 | 1.33, 3.25 | **0.001** |
| The other gender receiving more responsibilities at work: Yes (vs ref.: Neutral+No) | 618 | 2.21 | 1.25, 3.87 | **0.006** |
| The other gender getting more recognition for equal achievements: Yes (vs ref.: Neutral+No) | 618 | 1.92 | 1.17, 3.14 | **0.009** |

<sup>1</sup> OR = Odds Ratio, CI = Confidence Interval. Each variable was individually included in a logistic model adjusted for sex, age, Swiss language region and type of workplace.

### Sex-stratified analysis of factors associated with impaired mental well-being in physicians

As a next step, we hypothesized that some personal or workplace-related factors are important only in one sex, regarding their potential association with impaired mental well-being in physicians. We therefore used univariable and multivariable logistic regression models in the sex-stratified subgroups of the study populations. We have conducted univariable analyses in women (Table 5) and in men (Table 6), as well as multivariable analyses in women (Table 7) and in men (Table 8). Pregnancy-specific questions revealed a noteworthy finding that a failure to comply with Swiss labor laws regarding maximum working hours during a past or current pregnancy was strongly associated with currently having an impaired PWBI. This was apparent in both univariable (Table 5) and multivariable analysis (Table 7). Next, a further major sex difference among the sex-stratified analyses were that in women only - but not in men - a negative view on parenthood at the workplace was associated with a higher probability of impaired PWBI. By contrast, in men only, life partner’s support for their career was associated with PWBI (Table 8), whereas no evidence of such an association was present in women (Table 7). There was evidence in both sexes in univariable and/or adjusted analyses that the intention or history of delaying having children was associated with an impaired PWBI. Finally, gender-related discrimination was to a much larger extent associated with PWBI in women than in men. In summary, the sex-stratified analyses revealed some distinct properties how men and women among the GIM physician workforce may show different relationships between personal or workplace-related factors and their overall mental health, whereas the intention to delay having children tended to associate with impaired PWBI.

**Table 5.** Univariable logistic models for impaired physician well-being index in women.

|  |  |  |  |  |
| --- | --- | --- | --- | --- |
| **Characteristic** | **N** | **OR<sup>1</sup>** | **95% CI<sup>1</sup>** | **p-value** |
| Having a good network: Yes (vs ref.: No) | 347 | 0.45 | 0.26, 0.78 | **0.005** |
| Being unhappy with work: Yes (vs ref.: No) | 364 | 9.18 | 5.09, 16.8 | **<0.001** |
| Number of children (per child) | 392 | 0.96 | 0.72, 1.24 | 0.77 |
| Having good mentoring: Yes (vs ref.: No) | 347 | 0.52 | 0.30, 0.91 | **0.024** |
| Compliance with labor laws regarding working hours during pregnancy: No (vs ref.: Yes) | 131 | 4.99 | 1.57, 22.3 | **0.014** |
| Delaying having children (ref.: Have not delayed) | 274 |  |  |  |
| have delayed |  | 2.45 | 0.98, 6.39 | 0.058 |
| will delay |  | 2.01 | 0.90, 4.87 | 0.1 |
| will not delay |  | 1.75 | 0.63, 4.88 | 0.28 |
| Having partner's support for professional advancement: No (vs ref.: Neutral+Yes) | 294 | 1.42 | 0.31, 4.84 | 0.6 |
| My workplace has a culture valuing contributions of both women and men: No (vs ref.: Neutral+Yes) | 376 | 2.24 | 1.11, 4.34 | **0.02** |
| At my workplace, people of different origin are respected: No (vs ref.: Neutral+Yes) | 376 | 2.41 | 0.88, 6.13 | 0.072 |
| At my workplace, a safe environment is promoted in which different people can express their true self: No (vs ref.: Neutral+Yes) | 376 | 2.64 | 1.51, 4.58 | **<0.001** |
| Interactions between people of different groups (e.g gender, ethnicity, age) are trustful and respectful at my workplace: No (vs ref.: Neutral+Yes) | 376 | 2.25 | 0.88, 5.35 | 0.074 |
| **Gender discrimination by... / Because of my gender ...** |  |  |  |  |
| Not being taken seriously: Yes (vs ref.: Neutral+No) | 370 | 1.71 | 1.01, 2.89 | **0.046** |
| Others having less confidence in me: Yes (vs ref.: Neutral+No) | 370 | 1.13 | 0.65, 1.94 | 0.66 |
| Feeling discriminated at work: Yes (vs ref.: Neutral+No) | 370 | 2 | 1.13, 3.48 | **0.016** |
| Being treated worse by colleagues: Yes (vs ref.: Neutral+No) | 370 | 4.37 | 2.48, 7.72 | **<0.001** |
| Smaller perceived chance to rise in hierarchy: Yes (ref.: Neutral+No) | 370 | 1.37 | 0.81, 2.31 | 0.24 |
| Often taking on duties that don't advance one's career: Yes (vs ref.: Neutral+No) | 370 | 1.63 | 0.97, 2.79 | 0.069 |
| The other gender receiving more responsibilities at work: Yes (vs ref.: Neutral+No) | 370 | 2.25 | 1.28, 3.93 | **0.004** |
| The other gender getting more recognition for equal achievements: Yes (vs ref.: Neutral+No) | 370 | 1.99 | 1.18, 3.37 | **0.01** |
| How parenthood is viewed at the workplace: Negative (ref.: Neutral+Good) | 114 | 2 | 0.50, 6.72 | 0.28 |
| <sup>1</sup> OR = Odds Ratio, CI = Confidence Interval. Each variable was individually included in an univariable logistic model. |  |  |  |  |

**Table 6.**
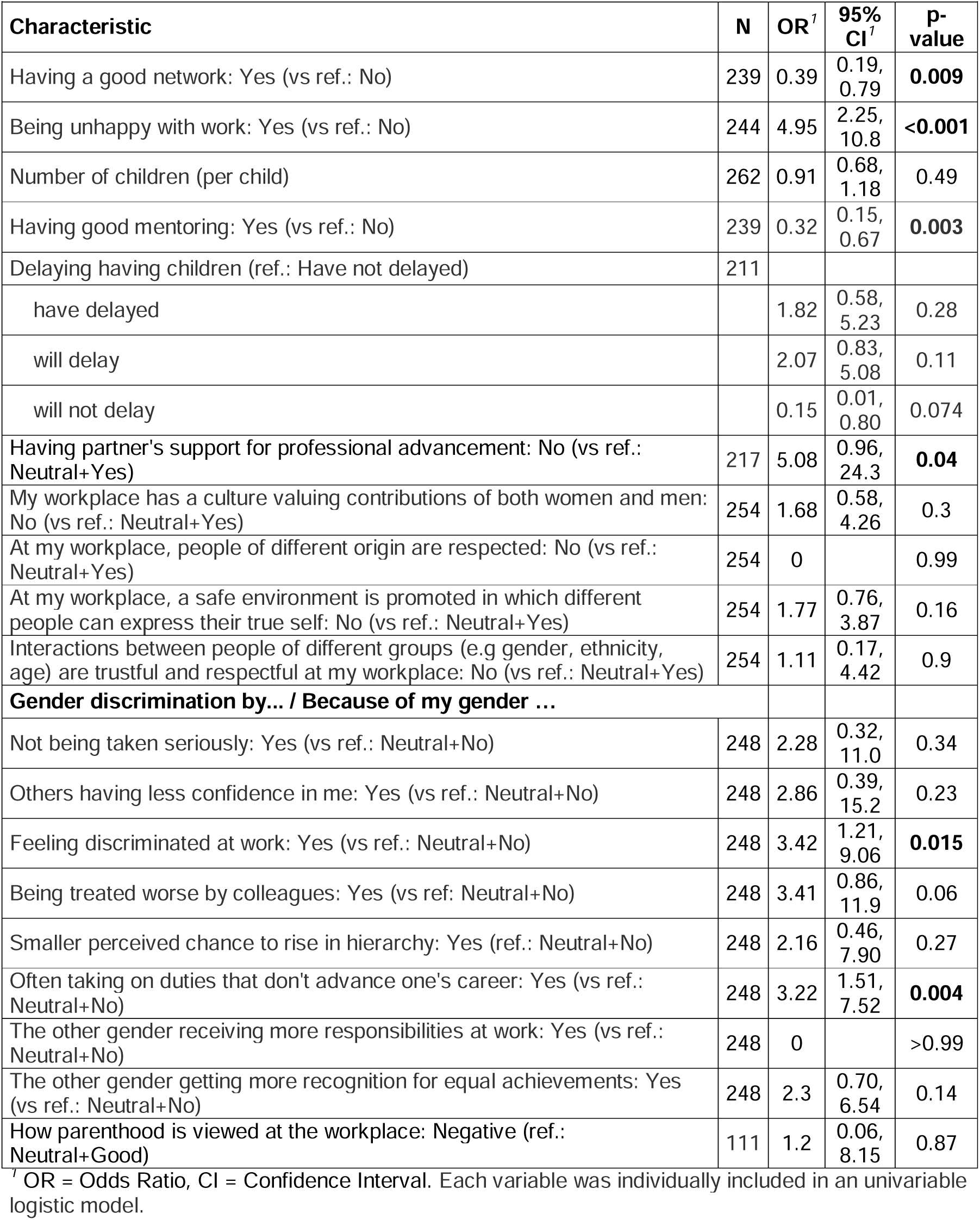
Univariable logistic models for impaired physician well-being index in men.

**Table 7.**
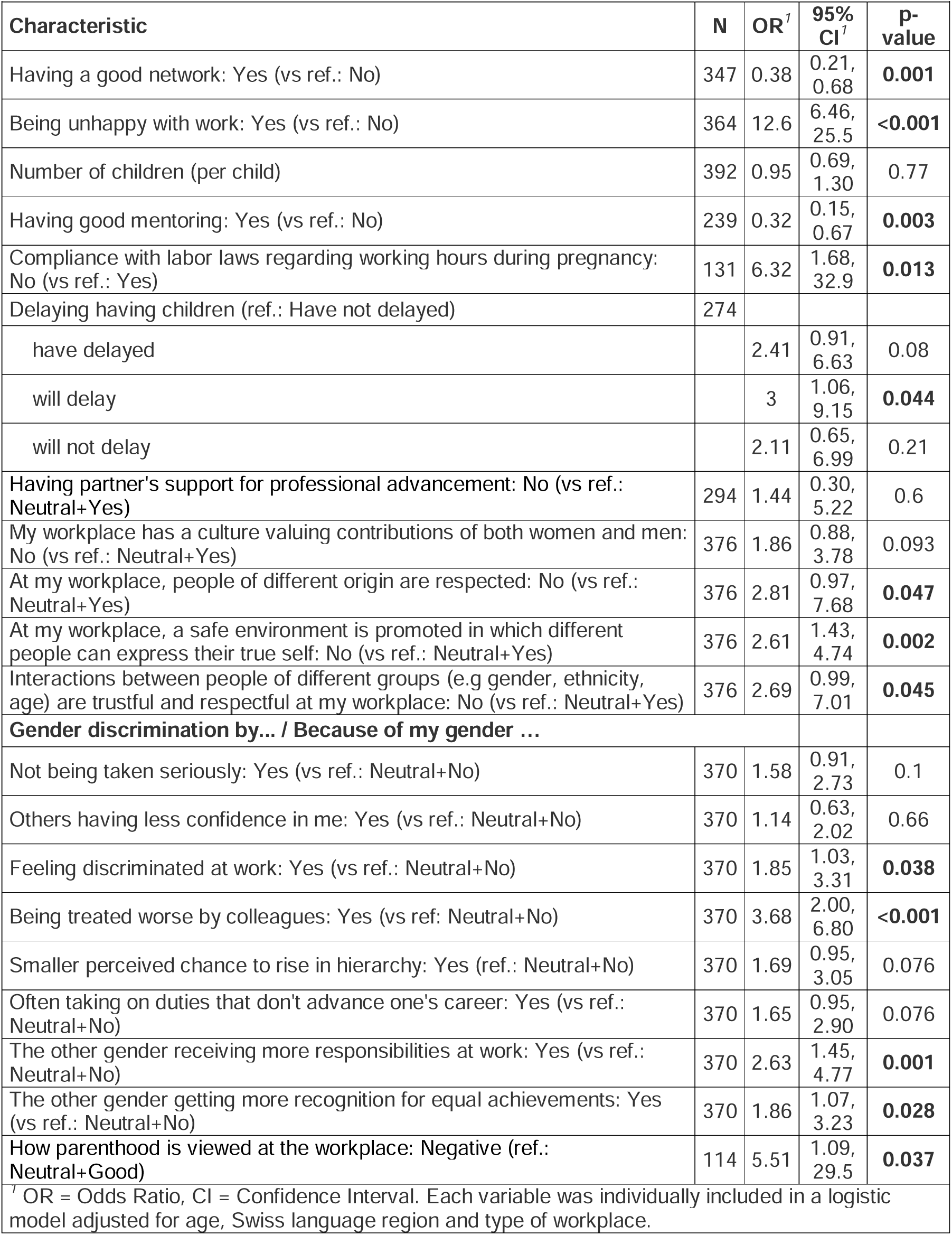
Multivariable logistic models for impaired physician well-being index in women.

|  |  |  |  |  |
| --- | --- | --- | --- | --- |
| **Characteristic** | **N** | **OR<sup>1</sup>** | **95% CI<sup>1</sup>** | **p-value** |
| Having a good network: Yes (vs ref.: No) | 347 | 0.38 | 0.21, 0.68 | **0.001** |
| Being unhappy with work: Yes (vs ref.: No) | 364 | 12.6 | 6.46, 25.5 | **<0.001** |
| Number of children (per child) | 392 | 0.95 | 0.69, 1.30 | 0.77 |
| Having good mentoring: Yes (vs ref.: No) | 239 | 0.32 | 0.15, 0.67 | **0.003** |
| Compliance with labor laws regarding working hours during pregnancy: No (vs ref.: Yes) | 131 | 6.32 | 1.68, 32.9 | **0.013** |
| Delaying having children (ref.: Have not delayed) | 274 |  |  |  |
| have delayed |  | 2.41 | 0.91, 6.63 | 0.08 |
| will delay |  | 3 | 1.06, 9.15 | **0.044** |
| will not delay |  | 2.11 | 0.65, 6.99 | 0.21 |
| Having partner's support for professional advancement: No (vs ref.: Neutral+Yes) | 294 | 1.44 | 0.30, 5.22 | 0.6 |
| My workplace has a culture valuing contributions of both women and men: No (vs ref.: Neutral+Yes) | 376 | 1.86 | 0.88, 3.78 | 0.093 |
| At my workplace, people of different origin are respected: No (vs ref.: Neutral+Yes) | 376 | 2.81 | 0.97, 7.68 | **0.047** |
| At my workplace, a safe environment is promoted in which different people can express their true self: No (vs ref.: Neutral+Yes) | 376 | 2.61 | 1.43, 4.74 | **0.002** |
| Interactions between people of different groups (e.g gender, ethnicity, age) are trustful and respectful at my workplace: No (vs ref.: Neutral+Yes) | 376 | 2.69 | 0.99, 7.01 | **0.045** |
| **Gender discrimination by... / Because of my gender ...** |  |  |  |  |
| Not being taken seriously: Yes (vs ref.: Neutral+No) | 370 | 1.58 | 0.91, 2.73 | 0.1 |
| Others having less confidence in me: Yes (vs ref.: Neutral+No) | 370 | 1.14 | 0.63, 2.02 | 0.66 |
| Feeling discriminated at work: Yes (vs ref.: Neutral+No) | 370 | 1.85 | 1.03, 3.31 | **0.038** |
| Being treated worse by colleagues: Yes (vs ref.: Neutral+No) | 370 | 3.68 | 2.00, 6.80 | **<0.001** |
| Smaller perceived chance to rise in hierarchy: Yes (ref.: Neutral+No) | 370 | 1.69 | 0.95, 3.05 | 0.076 |
| Often taking on duties that don't advance one's career: Yes (vs ref.: Neutral+No) | 370 | 1.65 | 0.95, 2.90 | 0.076 |
| The other gender receiving more responsibilities at work: Yes (vs ref.: Neutral+No) | 370 | 2.63 | 1.45, 4.77 | **0.001** |
| The other gender getting more recognition for equal achievements: Yes (vs ref.: Neutral+No) | 370 | 1.86 | 1.07, 3.23 | **0.028** |
| How parenthood is viewed at the workplace: Negative (ref.: Neutral+Good) | 114 | 5.51 | 1.09, 29.5 | **0.037** |
| <sup>1</sup> OR = Odds Ratio, CI = Confidence Interval. Each variable was individually included in a logistic model adjusted for age, Swiss language region and type of workplace. |  |  |  |  |

**Table 8.** Multivariable logistic models for impaired physician well-being index in men.

|  |  |  |  |  |
| --- | --- | --- | --- | --- |
| **Characteristic** | **N** | **OR<sup>1</sup>** | **95% CI<sup>1</sup>** | **p-value** |
| Having a good network: Yes (vs ref.: No) | 239 | 0.35 | 0.16, 0.77 | **0.009** |
| Being unhappy with work: Yes (vs ref.: No) | 244 | 4.98 | 2.18, 11.4 | **<0.001** |
| Number of children (per child) | 262 | — | — |  |
| Having good mentoring: Yes (vs ref.: No) | 239 | 0.25 | 0.11, 0.55 | **<0.001** |
| Delaying having children (ref.: Have not delayed) | 211 |  |  |  |
| have delayed |  | 1.82 | 0.55, 5.59 | 0.3 |
| will delay |  | 1.39 | 0.45, 4.33 | 0.57 |
| will not delay |  | 0.07 | 0.00, 0.47 | **0.022** |
| Having partner's support for professional advancement: No (vs ref.: Neutral+Yes) | 217 | 5.93 | 1.06, 30.9 | **0.031** |
| My workplace has a culture valuing contributions of both women and men: No (vs ref.: Neutral+Yes) | 254 | 1.38 | 0.46, 3.69 | 0.54 |
| At my workplace, people of different origin are respected: No (vs ref.: Neutral+Yes) | 254 | 0 |  | 0.99 |
| At my workplace, a safe environment is promoted in which different people can express their true self: No (vs ref.: Neutral+Yes) | 254 | 1.55 | 0.64, 3.55 | 0.31 |
| Interactions between people of different groups (e.g gender, ethnicity, age) are trustful and respectful at my workplace: No (vs ref.: Neutral+Yes) | 254 | 0.98 | 0.14, 4.17 | 0.98 |
| **Gender discrimination by... / Because of my gender ...** |  |  |  |  |
| Not being taken seriously: Yes (vs ref.: Neutral+No) | 248 | 2.03 | 0.28, 10.1 | 0.42 |
| Others having less confidence in me: Yes (vs ref.: Neutral+No) | 248 | 2.86 | 0.37, 16.9 | 0.26 |
| Feeling discriminated at work: Yes (vs ref.: Neutral+No) | 248 | 3.24 | 1.12, 8.86 | **0.024** |
| Being treated worse by colleagues: Yes (vs ref.: Neutral+No) | 248 | 3.19 | 0.77, 11.7 | 0.087 |
| Smaller perceived chance to rise in hierarchy: Yes (ref.: Neutral+No) | 248 | 1.76 | 0.36, 6.69 | 0.43 |
| Often taking on duties that don't advance one's career: Yes (vs ref.: Neutral+No) | 248 | 3.29 | 1.51, 7.81 | **0.004** |
| The other gender receiving more responsibilities at work: Yes (vs ref.: Neutral+No) | 248 | 0 |  | 0.99 |
| The other gender getting more recognition for equal achievements: Yes (vs ref.: Neutral+No) | 248 | 2.07 | 0.61, 6.10 | 0.21 |
| How parenthood is viewed at the workplace: Negative (ref.: Neutral+Good) | 111 | 1.37 | 0.06, 11.0 | 0.79 |

<sup>1</sup> OR = Odds Ratio, CI = Confidence Interval. Each variable was individually included in a logistic model adjusted for age, Swiss language region and type of workplace.

## Discussion

### Statement of principal findings

The principal findings of this study are that a considerable fraction of participating GIM physicians in Switzerland showed an impaired well-being, with some evidence of a higher probability in women than in men. We found congruent association between many personal or workplace-related factors and PWBI, including workplace inclusiveness, and mentoring and having a good professional network. However, in women gender-related discrimination and perceiving a negative view of parenthood at the workplace were associated with PWBI, whereas in men rather support of one’s career by the spouse was associated with PWBI. One of the strongest predictor of impaired PWBI among physician-mothers was incompliance with labor laws during pregnancy. Overall, when assuming some degree of causality in these associations, there are important sociocultural mechanisms contributing to physician well-being that affect women and men differently among GIM physicians in Switzerland.

### Strengths and weaknesses of the study

A novelty and strength of this study is the profound sex-stratified assessment of personal and workplace factors associated with physician well-being, which allows unprecedented insights in the Swiss physician workforce compared to previous work (5,11,22–24). Further, a wide diversity of GIM physicians is represented in this dataset, including all ages, respondents of all Swiss language regions and different types of workplaces. Many previous studies used less diverse samples of only GPs or only trainees (5,11,22). Next, a strength is the use and cross-validation of a survey in the two most spoken languages of Switzerland. Another strength is the overall consistency in finding of impaired well-being in physicians in comparison to similar studies in the past: Lindemann et al., Zumbrunn et al., Sebo et al. and most recently Villiger et al. all showed a prevalence of 20-33% of impaired well-being in different populations of physicians in Switzerland (5,11,22–24). This study also has some weaknesses. First, we were unable to calculate a response rate for this survey due to the lack of a precisely defined sampling frame with use of the physicians’ society newsletter e- mail and a journal-advertised survey participation. However, two previous surveys in similar populations of physicians in Switzerland both yielded at least 40% response rates (5,11), while enrolling a very similar fraction of physicians with impaired PWBI. Further, for the number of associations tested in this analysis, no adjustments were made for multiple testing. Finally, the current cross-sectional study design makes it impossible to determine causality of any reported associations.

### Meaning of the study: possible mechanisms and implications for clinicians or policymakers

The present data reveal an impaired well-being in many Swiss physicians that remind of U.S. data, where especially female physicians are more likely had to delay personal life decisions, face gender-related discrimination and obstacles in their professional development, all of which may contribute to burnout (25,26). Based on the observed findings, consequently a combination of both general and sex-specific approaches could be considered to tackle impaired physician well-being in women and men, assuming that the obtained associations were the result from some underlying degree of causality and no predominant reverse causality. Potential common starting points to generally improve physician well-being among both sexes may include improving a workplace climate that is inclusive for all and excludes no minorities, regardless of gender, sexual orientation, ethnicity or religion. While this may seem obvious, in medicine this lack of inclusivity can still be considered a prevalent problem that is going to require further attention in the coming years, with the increasing fraction of female physicians (27,28). In addition, also reducing burnout tendencies by diminishing administrative workloads has been suggested as a promising strategy to improve physician well-being by surveys both internationally and in Switzerland (2,3,24,29,30). Finally, we found that the well-being of men among the participating physicians tends to be sensitive towards a lack of career support by their spouse, whereas women among the participating physicians predominantly showed impaired well-being in association with gender-related discrimination, but no association with spouses’ support. Because the population structure of male and female physicians slightly differed, e.g. more older colleagues were included versus more participants were female among the younger participants. Therefore, the multivariable analyses with adjustments for age were crucial, because younger physicians may simply not have a long-time life partner yet that may have a say on their career. As the finding was robust after adjustment, this indicates that among male physicians a traditional gender role may persist in some way, as their workplace environment potentially expects them to not take care of children or household in a way that it affects their career or availability for shiftwork or full-time employment. Thus, we speculate that in male physicians it is usually their spouse that takes care of children and misses work when children are sick, whereas these tasks are rather not the role of the spouse of female physicians, but a task covered by the female physicians themselves, as has been shown to be the case for male and female employees among the general population in Switzerland (31,32). This sociocultural norm, or traditional gender norm, may be something worth investigating further, as it may be the root of some significant obstacles for the well-being of physicians: In a more ideal world, male physicians should not be expected to carry forward the traditional male gender role at work, whereas female physicians should not be in a constant conflict between fulfilling professional duties, resisting discrimination and poor inclusivity, yet carry the majority of mental and/or physical load of housework and childcare at the same time. For this conflict, it would be of utmost importance to improve the quality and affordability of external childcare possibilities in Switzerland, to derive strategies for what to do when employees are sick, or to better enforce that labor laws are respected during pregnancy.

### Unanswered questions and future research

Several efforts to improve physician well-being have been initiated in different countries (33–35). However, how effective targeted interventions are in improving physician well-being is not well established. As suggested by some, an adaptation of personal character traits such as resilience and positivity to cope with clinical workloads and demands may help some individuals (36) but is not a promising long-term solution towards improving physician well-being in an underfunded and understaffed healthcare environment. A particular type of intervention to be evaluated that may improve not only some patients’ healthcare participation but also physician well-being, could be telemedicine, the practice of which has been associated with an improved well-being among Swiss physicians in an emerging report (37). We further see opportunities for innovation towards improving physician well-being, including gender-adapted interventions that may involve flexible working arrangements, telemedicine or childcare-adapted shift schedules, but also inclusivity and co-worker valorization by institutions and stakeholders in medicine.

### Conclusions and recommendations

The present data reveal that among GIM physicians in Switzerland, the challenges associated with impaired physician well-being overlap only in part between women and men. Features more likely to be associated with impaired physician well-being in women physicians include gender-related discrimination and among physician-mothers the incompliance with labor laws during pregnancy, whereas in men physicians a lack of career support by their spouse was associated with impaired physician well-being – reflecting in some extent the traditional gender role models present in medicine. To improve the well-being of healthcare professionals, considerable policy efforts and incentives are needed that are tailored for each sex. Such efforts may not come cost-neutral in the short-term, but they may be an investment with a good return in the long run, improving not only physician health but also preventing attrition from the physician workforce.

## Acknowledgements

The authors are thankful to the respondents, and to the support by Swiss Society of Internal Medicine, Swiss Young General Practitioners Association (JHaS), Primary Hospital Care and the hospitals involved with distribution of the survey.

## Data availability

All data produced in the present study are available upon reasonable request to the authors.

